# Role of demographic factors and vascular anatomy in successful trans-radial Tiger catheterization in coronary angiography

**DOI:** 10.1101/2023.02.03.23285432

**Authors:** Ali Asghar Farsavian, Erfan Ghadirzadeh, Maryam Daghestani, Samad Golshani, Ali Asghar Nadi Ghara

## Abstract

**Objective:** The use of a single catheter in the trans-radial angiography approach (TRA) is associated with fewer complications. However, there are some discrepancies regarding the effectiveness of these catheters. Catheter selection is crucial in reducing the risk of trauma to the coronary arteries in angiography, so the main objective of the present study was to investigate the success rate of the Tiger single catheter in the right and left coronary angiography in the TRA, in addition to characterizing the role of demographic factors, underlying conditions and vascular anatomy in its success.

**Methods:** This observational study was conducted on 127 patients with suspected CAD. Demographic data and risk factors were recorded in a checklist. The root diameter of the aortic artery at different sections ejection fraction was measured by transthoracic echocardiography before performing TRA. Coronary anomaly, tortuosity of the radial artery pathway to the aorta, as well as coronary artery take-off and procedure indication, were also recorded during the TRA.

**Results:** In this study, 127 patients with a mean age of 59 ± 10.3 were enrolled. Sixty-seven patients (51.5%) were female, and 63 patients (48.5%) were male. Failure was reported only in 5 patients (3.85%). Age, gender, BMI, height, aortic diameter, coronary anomalies, tortuosity, hypertension, dyslipidemia, and smoking had not a significant relationship with successful Tiger catheterization (P > 0.05) but high coronary take-off had (P= 0.004).

**Conclusion:** In cases of high coronary take-off and coronary abnormality, a Tiger catheter can probably benefit the patient with a trans-radial approach.

## Introduction

Coronary artery diseases (CAD) are important causes of mortality in the world and one of the most critical priorities related to the health care system (1). Various diagnostic methods assist the physician in diagnosis and therapeutic decision-making in CAD. Among them, cardiovascular angiography is one of the most important and accurate cardiac tests used nowadays (2).

Coronary angiography can be performed with a catheter via radial or femoral approaches. The trans-radial approach (TRA) is performed through the radial artery and has been claimed to impose fewer restrictions on the patient than femoral angiography (3). Catheter selection is crucial to the quality of angiography, as it reduces the risk of coronary artery trauma and offers an appropriate platform for instrument transfer (4). The Judkins catheter is the most commonly used catheter in coronary angiography. Nonetheless, it has limitations and disadvantages when utilized in a TRA (5, 6). To properly conduct a TRA, both the right and left coronary arteries must be engaged. Consequently, the use of a Judkins catheter would involve the use of both a right and a left Judkins catheter (5, 6). Nevertheless, in order to do that, a catheter exchange must happen during the procedure. Exchanging catheters during a TRA would cause undesirable complications. For instance, radial artery spasms may occur during catheter exchange and lead to catheter entrapment and even subclavian artery dissection in some cases (5-7). New catheters, such as the Tiger Radial TIG, were developed to overcome such limitations. These catheters have primary and secondary curves, which theoretically allow the interventional cardiologist to use a single catheter for TRA (5). This leads to fewer complications, less contrast administration, and shorter fluoroscopy time (8-10). However, multiple studies that investigated the success rate of the Tiger single catheter reported values ranging from 50 to 90 percent, and sometimes failure, with no significant difference in the mean contrast administration, complication rate, or fluoroscopy time in comparison to the Judkins catheter (4, 8, 11, 12).

Since there have been inconsistencies in the results of previous research, and since the evaluation of the relationship between coronary anatomy and the selected Tiger catheter has not been studied in the Iranian population, the main objective of the present study was to investigate the success rate of the Tiger single catheter in the right and left coronary angiography in the trans-radial approach (TRA), in addition to characterizing the role of demographic factors associated with the success and failure of this catheter.

## Materials and Methods

This observational study was conducted on 127 patients with suspected CAD who were candidates for trans-radial coronary angiography at the Mazandaran Heart Center, Sari, Iran. The study size was calculated based on Langer et al. (13) results. All CAD elective candidates for angiography were enrolled. Exclusion criteria were hypotension (systolic blood pressure less than 100), inability to touch the radial artery pulse or a positive Allen test, inability to execute TRA for any reason or any contraindications other than catheter engagement, and refusal to continue participating in the study. This research was reviewed and approved by the research ethics committee of Mazandaran University of Medical Sciences (Ethics approval number IR.MAZUMS.REC.1398.5497), and written informed consent was obtained from the participants to include the clinical details. All procedures adhered to the Declaration of Helsinki’s ethical guidelines. No intervention was performed on the patients by the research team, and all information was collected from medical records, angiography reports, and observation.

Demographic data, including age, gender, height, body mass index (BMI), and risk factors such as diabetes mellitus (DM), dyslipidemia (DLP), and hypertension (HTN), were recorded in a checklist. Blood samples were taken from patients to measure the levels of triglycerides (TG), total cholesterol (CHOL), high-density lipoproteins (HDL), and low-density lipoproteins (LDL), as well as fasting blood sugar (FBS). Before performing TRA, the root diameter of the aorta artery at different sections and ejection fraction (EF) were measured using transthoracic echocardiography. Coronary anomaly, tortuosity of the radial artery to aorta pathway, coronary artery take-off, and procedure indication were also documented during the TRA. Before performing the procedure, the Allen test was used to assess the adequacy of ulnar artery blood flow. Local anesthesia was injected subcutaneously with a 25-gauge needle. The radial artery was punctured using a 20-gauge needle. 4F, 5F, or 6F arterial sheets were used. After reaching the arterial route, intravenous verapamil or nitroglycerine was administered by the physician, and then the Tiger single catheter was used to execute the TRA. Failure of the TRA leads to the change of vascular direction to the opposite radial or femoral artery, and it was defined as follows: inability to access the radial artery due to unsuccessful arterial puncture, inability to engage the catheter properly, or replacement of the Tiger catheter with other catheters such as the right and left Judkins. After completing the procedure, all instruments were withdrawn, and the TR Band homeostasis was embedded and inflated for the Arteriotomy site. The sheet was removed from the lip coat before the patient’s departure, and then the TR Band volume was emptied constantly within two hours.

First, normality was assessed using the Shapiro-Wilk test, and if the distribution was normal, an ANOVA analysis, an independent samples T-test, and a chi-square test were performed. However, if the data were not normally distributed, non-parametric equivalent tests such as chi-square, Mann-Whitney U, and Kruskal-Wallis tests were used. After data collection, data analysis was performed using SPSS software version 24 and STATA software version 14. A P value of less than 0.05 was considered statistically significant.

## Results

In total, 127 patients were enrolled. The success rate in left and right angiography with the Tiger single catheter was 96.15%. Only one case (0.78%) exhibited severe spasms while using the Tiger catheter. Patients in the “successful” group had a mean age of 57.99 ± 10.40 years, while those in the “unsuccessful” group had a mean age of 61.60 ± 9.9 years. Patients were divided into two categories, under 50 and over 50 years old. Among people with successful angiography, 23.2% of the patients were under 50, and 76.8% were over 50 years old. 49.6% of the patients were men. In the unsuccessful group, the mean height was 193.80 cm, whereas the successful group’s mean height was 165.09 cm. The mean BMI in the unsuccessful group was 26.81, while the mean BMI in the successful group was 27.43. We discovered no statistically significant relationship between successful Tiger angiography and age, gender, height, or BMI (P = 0.868, 0.366, 0.709, and 0.728, respectively) (Table 1). Our analysis also indicates no significant relationship between successful Tiger catheterization and HTN, DLP, DM, or smoking. (P = 0.887, 0.971, 0.915, and 0.193, respectively) (Table 1). Blood indices were also measured in the patients. None of the parameters were found to be significantly associated with a successful Tiger angiogram (P > 0.05) (Table 2). EF was measured by transthoracic echocardiography. No significant relationship was found between EF and Tiger TRA succession (P = 0.938) (Table 2).

**Table 1.** The relationship between successful Tiger catheterization in TRA and demographic and risk factors.

| Variable |  | Unsuccessful | Successful | P value<br>(P) | Odds<br>Ratio<br>(OR) | 95% Confidence |  |
| --- | --- | --- | --- | --- | --- | --- | --- |
|  |  |  |  |  |  | Interval (CI) |  |
|  |  |  |  |  |  | Lower | Upper |
|  |  | Number<br>(Percent) | Number<br>(Percent) |  |  |  |  |
| Age | Under 50 | 1 (20%) | 29 (23.2%) | 0.868 | 0.828 | 0.089 | 7.699 |
|  | Over 50 | 4 (80%) | 69 (76.8%) |  |  |  |  |
| Gender | Female | 4 (80%) | 63 (50.4%) | 0.366 | 3.937 | 0.428 | 36.215 |
|  | Male | 1 (20%) | 62 (49.6%) |  |  |  |  |
| HTN | YES | 2 (40%) | 54 (43.2%) | 0.887 | 0.887 | 0.141 | 5.431 |
|  | NO | 3 (60%) | 71 (56.8%) |  |  |  |  |
| DLP | YES | 3 (60%) | 49 (39.2%) | 0.971 | 1.034 | 0.167 | 6.413 |
|  | NO | 2 (40%) | 76 (60.8%) |  |  |  |  |
| DM | YES | 3 (60%) | 72 (57.6%) | 0.915 | 1.104 | 0.178 | 6.642 |
|  | NO | 2 (40%) | 53 (42.4%) |  |  |  |  |
| Smoking | YES | 5 (100%) | 93 (74.4%) | 0.193 | 0.949 | 0.906 | 0.994 |
|  | NO | 0 (0%) | 32 (25.6%) |  |  |  |  |

|  |  | Mean ± SD | Mean ± SD |  |  |  |  |
| --- | --- | --- | --- | --- | --- | --- | --- |
| Height |  | 163.80 ± 9.47 | 165.09 ± 7.59 | 0.709 | 1.023 | 0.909 | 1.151 |
| BMI |  | 26.81 ± 2.96 | 27.43 ± 3.96 | 0.728 | 1.044 | 0.819 | 1.331 |

**Table 2.** Evaluation of blood indices and EF in two angiographic groups.

| Variable | Unsuccessful |  | Successful |  | P | OR | 95% CI |  |
| --- | --- | --- | --- | --- | --- | --- | --- | --- |
|  | Mean | SD | Mean | SD |  |  | Lower | Upper |
| EF | 55 | 3.54 | 52.38 | 7.39 | 0.430 | 0.938 | 0.799 | 1.100 |
| FBS | 114.80 | 26.20 | 112.01 | 48.62 | 0.898 | 0.999 | 0.982 | 1.016 |
| TG | 177.20 | 50.61 | 146.79 | 89.75 | 0.459 | 0.997 | 0.990 | 1.005 |
| COL | 207.40 | 31.72 | 158.92 | 52.16 | 0.053 | 0.986 | 0.972 | 1.000 |
| HDL | 44.20 | 4.97 | 38.29 | 8.26 | 0.118 | 0.921 | 0.831 | 1.021 |
| LDL | 115 | 26.73 | 93.93 | 41.08 | 0.265 | 0.990 | 0.927 | 1.008 |

The relationship between “successful” Tiger angiography in TRA and aortic diameter in different sections was not statistically significant (P > 0.05) (Table 3) (Figure 1). In addition, the aortic diameters at different sections were categorized into two categories (normal and abnormal). According to the findings, 73 patients with an abnormal D3 status underwent a successful Tiger angiogram, while none experienced a failure. Only the relationship between successful Tiger angiography and abnormal D3 aortic diameter was statistically significant (P = 0.015) (Table 4).

**Table 3.**
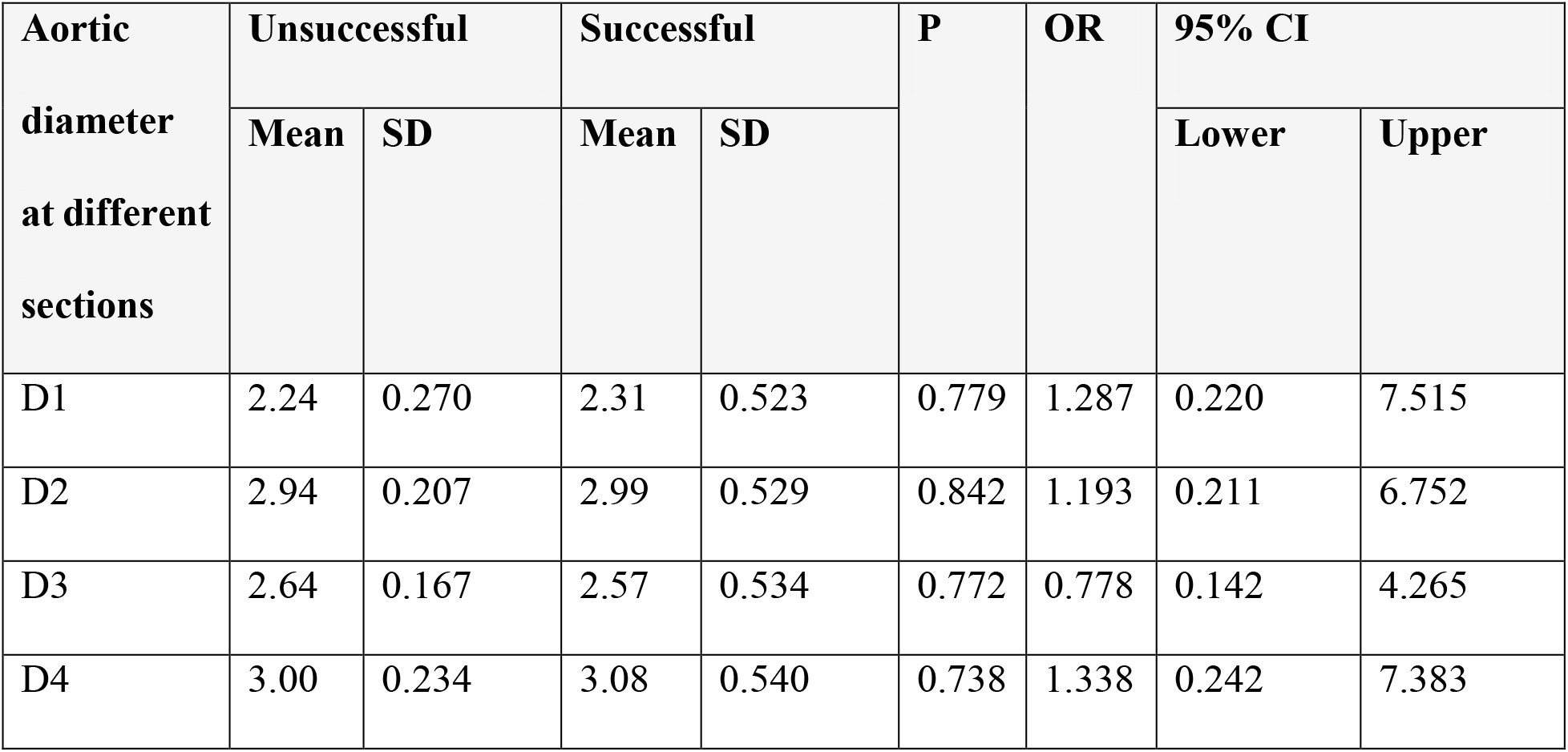
The relationship between successful Tiger single catheter angiography in left and right coronary arteries in trans-radial method, and aortic diameter in different sections

| Aortic diameter at different sections | Unsuccessful |  | Successful |  | P | OR | 95% CI |  |
| --- | --- | --- | --- | --- | --- | --- | --- | --- |
|  | Mean | SD | Mean | SD |  |  | Lower | Upper |
| D1 | 2.24 | 0.270 | 2.31 | 0.523 | 0.779 | 1.287 | 0.220 | 7.515 |
| D2 | 2.94 | 0.207 | 2.99 | 0.529 | 0.842 | 1.193 | 0.211 | 6.752 |
| D3 | 2.64 | 0.167 | 2.57 | 0.534 | 0.772 | 0.778 | 0.142 | 4.265 |
| D4 | 3.00 | 0.234 | 3.08 | 0.540 | 0.738 | 1.338 | 0.242 | 7.383 |

**Figure 1.**
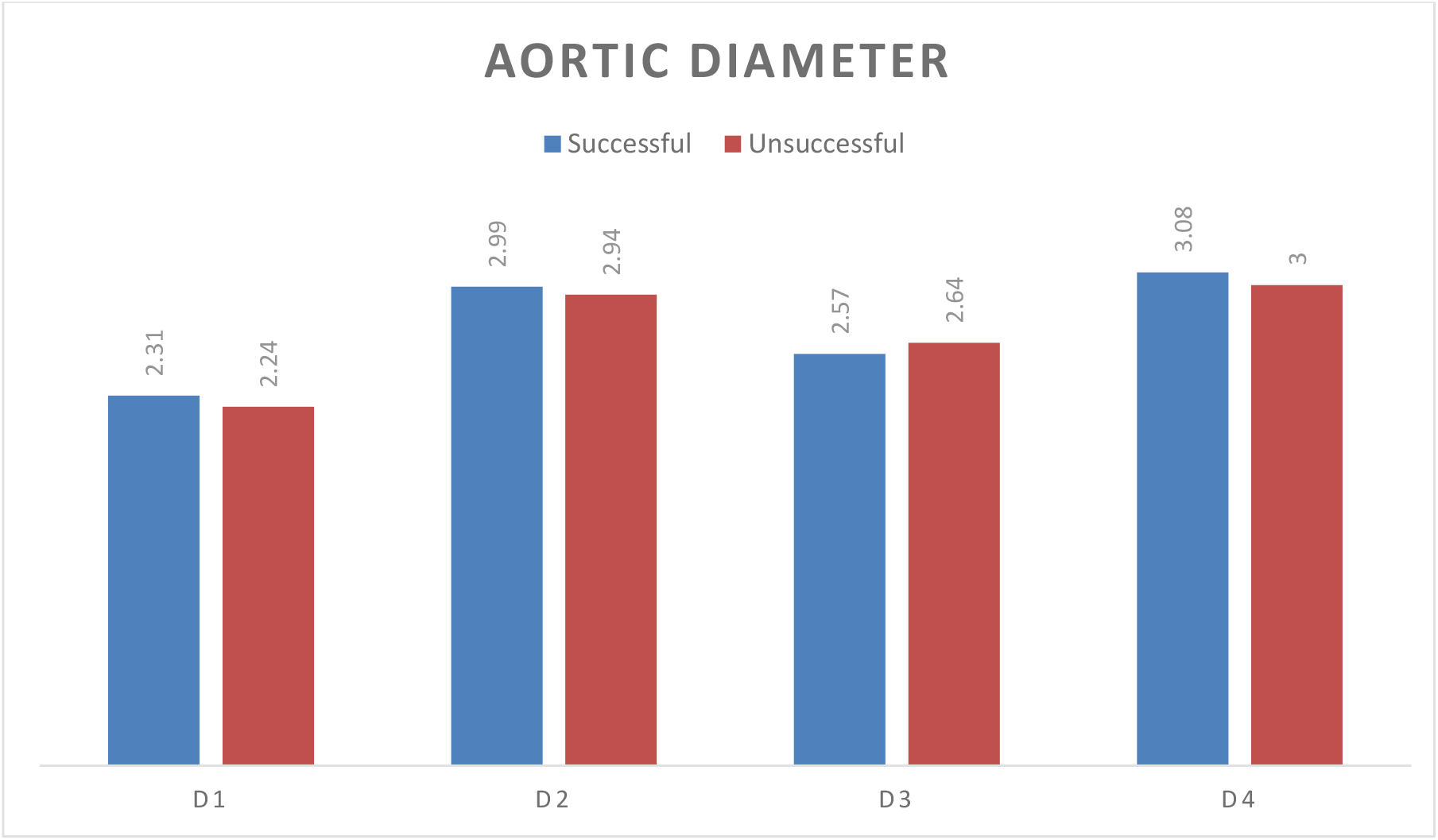
Mean of aortic diameter at different sections in both groups.

**Table 4.** Evaluation of aortic diameter indices in terms of normal and abnormal and its relationship with successful Tiger angiography.

| Aortic diameter |  | Unsuccessful |  | Successful |  | P | OR | 95% CI |  |
| --- | --- | --- | --- | --- | --- | --- | --- | --- | --- |
|  |  | Number | Percent | Number | Percent |  |  | Lower | Upper |
| D1 | Abnormal | 2 | 40 | 73 | 58.4 | 0.650 | 0.475 | 0.077 | 2.943 |
|  | Normal | 3 | 60 | 52 | 41.6 |  |  |  |  |
| D2 | Abnormal | 1 | 20 | 75 | 60 | 0.160 | 0.167 | 0.018 | 1.535 |
|  | Normal | 4 | 80 | 50 | 40 |  |  |  |  |
| D3 | Abnormal | 0 | 0 | 73 | 58.4 | 0.015 | 1.096 | 1.011 | 1.188 |
|  | Normal | 5 | 100 | 52 | 41.6 |  |  |  |  |
| D4 | Abnormal | 1 | 20 | 53 | 42.4 | 0.319 | 0.340 | 0.037 | 3.126 |
|  | Normal | 4 | 80 | 72 | 57.6 |  |  |  |  |

Coronary take-off, coronary anomaly and tortuosity were also assessed during TRA. Our analysis showed a statistically significant relationship between Tiger catheter succession and coronary take-off (P = 0.004) (Table 5). However, no significant relationship between the success of Tiger single catheter angiography and coronary anomalies or tortuosity was found (P = 0.776 and 0.447, respectively) (Table 5).

**Table 5.** The relationship between Tiger catheter succession in TRA and coronary tortuosity, anomaly, and take-off.

| Variable |  | Unsuccessful | Successful | P | OR | 95% CI |  |
| --- | --- | --- | --- | --- | --- | --- | --- |
|  |  |  |  |  |  | Lower | Upper |
|  |  | Number<br>(Percent) | Number<br>(Percent) |  |  |  |  |
| Tortuosity | NO | 5 (100%) | 112 (89.6%) | 0.447 | 0.957 | 0.921 | 0.995 |
|  | YES | 0 (0%) | 13 (10.4%) |  |  |  |  |
| Anomaly | NO | 5 (100%) | 123 (98.4%) | 0.776 | 0.961 | 0.928 | 0.995 |
|  | YES | 0 (0%) | 2 (1.6%) |  |  |  |  |
| Take-off | NO | 3 (60%) | 124 (99.2%) | 0.004 | 0.012 | 0.001 | 0.173 |
|  | YES | 2 (40%) | 1 (0.8%) |  |  |  |  |

## Discussion

Radial angiography is currently more popular than femoral angiography. Also, the use of a single catheter in TRA is associated with fewer complications, less use of contrast material, and less fluoroscopic time. Catheter selection is crucial to the quality of angiography, as it reduces the risk of coronary artery trauma and offers an appropriate platform for instrument transfer. So the main objective of the present study was to investigate the success rate of the Tiger single catheter in the right and left coronary angiography in the TRA, in addition to characterizing the role of demographic factors associated with the success and failure of this catheter.

In this study, the success rate in left and right angiography with the Tiger single catheter was 96.15%. No statistically significant relationship was observed between the success of Tiger single catheter angiography and demographic factors, including age, gender, BMI, height, HTN, DM, and smoking. Additionally, there was no significant relationship between successful Tiger angiography and coronary tortuosity, blood indices, dose length product, coronary anomalies, and aortic diameter in different sections. However, statistically significant relationships were observed between successful Tiger angiography and coronary take-off and abnormal D3 aortic diameter (P= 0.004 and 0.015, respectively).

In a study by Xanthopoulom et al. (10), the Tiger group had a lower rate of catheter replacement or access site change (17.8%) than the Judkins group (21.3%). This difference lacked statistical significance. Nevertheless, the required contrast volume, the angiography fluoroscopy time, and the rate of severe spasm (2.8% in their Tiger group) were significantly lower in the Tiger group compared to the Judkins. The amount of opacification in the LAD, RCA, and LCX arteries in the Tiger group was also significantly better than Judkins. The findings of the Xanthopoulom study contradicted those of this study. In fact, the rate of catheter replacement or lack of access in our study while using the Tiger catheter was only 3.85%, and only one patient showed severe spasm (0.78%).

In a study by Cadogan et al. (12), 38.5% of patients in the Tiger group and 17.1% in the Judkins group required additional catheters. The average use of additional equipment in the Judkins group was significantly lower than that in the Tiger group. The total amounts of dose area product, fluoroscopic time, mean used contrast volume, and catheterization time were similar between the two groups. Radial spasm was observed in 3 cases, and no other complications were observed. The total average expense was significantly lower in the Judkins group. They concluded that the use of the Judkins catheter as the selective catheter in cardiology is better in terms of cost-benefit and requires less equipment than the Tiger catheter. Their results were inconsistent with the present study.

In a study by Shroff et al. (9), the procedure performed with the Tiger single catheter method was associated with lower contrast volume and shorter fluoroscopy time compared to the Judkins two-catheter group. Also, the rate of forearm hematoma was lower in the Tiger catheter group. This study showed that the use of the Tiger catheter as a single catheter in TRA is very effective in reducing the amount of contrast, fluoroscopy time, and the rate of complications. The results of the above study are consistent with the present study.

In a study by Dehghani et al. (11) on 2100 patients, vascular complications occurred in 1% of the patients, and TRA failure occurred in 4.7%. Causes of TRA failure in their study included inability to reach the catheter to the ascending aorta in 51% of patients, inadequate support of the catheter guide in 36% of patients, and failure to puncture the radial artery in 13%. In the multivariate analysis, age above 75 years, previous history of coronary artery bypass graft (CABG), and height were identified as independent predictors of TRA failure. However, in our study, the rate of catheter replacement or lack of access in the Tiger group was only 3.85%, which was very low, and the spasm occurred in only one case. According to the obtained results, the Tiger catheter is considered a selective catheter in TRA.

In a study by Abdelaal et al. (4), unsuccessful primary TRA occurred in 2.7% of patients. The causes of failure in primary TRA included chronic radial artery occlusion in 11% of the patients, previous history of CABG in 27% of the patients, and cardiogenic shock in 20% of the patients. Failure in crossover TRA was observed in 1.8% of the patients. The causes of failure in performing Crossover TRA were inappropriate puncture in 57% of the patients, radial artery spasm in 17% of the patients, radial loop in 13% of the patients, subclavian tortuosity in 7% of the patients, and an inadequate support catheter guide in 7% of the patients. Female gender, previous history of CABG, and cardiogenic shock were independent predictors of TRA failure. However, our results showed no relationship between gender and TRA success.

## Conclusion

The results of this study showed that the success rate of the Tiger single catheter in the right and left coronary angiography with the trans-radial approach is very high (95.15%) and has no statistically significant relationship with underlying diseases (diabetes, dyslipidemia, and hypertension), demographic factors (age, gender, height, BMI), tortuosity, aortic diameter, and coronary anomaly. However, it does have a significant relationship with coronary take-off. In cases of high coronary take-off and coronary abnormality, a Tiger catheter can probably benefit the patient with a trans-radial approach.

## Data Availability

The data are available with the corresponding author and can be reached on request.

## Availability of data and materials

The data are available with the corresponding author and can be reached on request.

## Acknowledgments

None.

## Author’s contribution statement

SG and AF contributed to the conception and design of the work. MD and AF collected the data. MD and AN contributed to the interpretation of the data and analysis. EG did the writing of the manuscript and prepared the final version in addition to lingual and scientific editing of the manuscript. SG and AF revised the manuscript and did the final editing. All authors reviewed the paper and approved the final version of the manuscript.

